# Behavioral Assessment Reliability in Clinical Phenotyping and Biomarker Research for Autism

**DOI:** 10.64898/2026.01.24.25343227

**Authors:** Enrique Perez-Benavides, Jueqi Wang, Zijian Chen, Stefen Beeler-Duden, Zachary Jackokes, John D. Van Horn, Michael C. Schatz, Kevin A. Pelphrey, Archana Venkataraman

## Abstract

Autism Spectrum Disorder standardized behavioral assessments provide quantitative measures of symptoms, yet their reliability and consistency have not been systematically evaluated. We present the first large-scale comparative analysis of four widely used assessments. We analyzed behavioral assessments across three autism cohorts using correlations, clustering, and diagnostic agreement analyses. We related behavioral variation to genetic and imaging data to evaluate biomarker associations. Sentence-level embeddings generated by large language models reveal substantial semantic overlap across instruments. Nonetheless, behavioral scores are weakly correlated (0.26 ± 0.21), and diagnostic classification shows only 65–80% agreement between tests. These patterns hold across three datasets comprising N = 1 954. None of the assessments show consistent associations with widely studied MRI or genetic biomarkers. These findings expose critical inconsistencies among widely used autism assessments and underscore the need for more reliable tools to support precision phenotyping, biomarker discovery, and individualized care. Rather than diminishing the utility of behavioral assessment in autism, the inconsistencies identified here highlight a critical opportunity to refine how behavioral phenotypes are defined and operationalized.

## I. Introduction

Autism Spectrum Disorder (ASD) affects an estimated 1 in 31 children in the United States [1] and represents a significant public health concern due to its impact on individuals, families, and healthcare systems [2]. ASD is characterized by deficits in social communication and interaction, coupled with restricted and repetitive patterns of behavior [3]. Given its heterogeneity, the success of next-generation therapies will depend on accurate and personalized clinical phenotyping that can reliably capture individual symptom profiles. However, current practice relies on standardized behavioral assessments administered through caregiver interviews and clinician observations, raising concerns about the consistency and reproducibility of phenotypic measurement across settings and populations.

Key behavioral assessments used to quantify autistic symptoms include the Autism Diagnostic Observation Schedule (ADOS) [4], the Autism Diagnostic Interview–Revised (ADI-R) [5], the Social Responsiveness Scale (SRS) [6], and the Social Communication Questionnaire (SCQ) [7]. The ADOS, ADI-R, and SRS are the most frequently used instruments in both research and clinical practice for defining autism phenotypes [2]. The ADOS and ADI-R serve as gold-standard diagnostic tools based on clinician observation and caregiver report, respectively [8]. The SRS provides a dimensional measure of social behavior widely employed in largescale biomarker and genetic studies [9]. Finally, the SCQ is often used as an initial screening instrument. Implicit in the widespread use of these instruments is the assumption that–despite differences in format, content, and scoring–they provide reliable and congruent characterizations of autism. Although studies have evaluated them in large and diverse samples, challenges remain regarding their robustness and consistency when used more broadly at the population level.

This measurement uncertainty has far-reaching implications for ASD research, particularly as more studies seek to link behavioral variation with neuroimaging, genetics, and other highdimensional modalities. Behavioral assessments, especially the ADOS and ADI-R, are commonly treated as anchors for quantifying symptom severity and defining clinical subtypes. When these tools yield inconsistent classifications or severity scores, the resulting phenotypes may reflect methodological artifacts or subjective bias rather than true biological variation. This undermines the interpretability of reported associations and raises concerns about the validity of phenotype-driven subtyping in ASD. The issue is especially pressing in the context of large-scale precision psychiatry initiatives, which increasingly rely on behavioral assessments to delineate phenotypes for genetic and neurobiological analyses [9]. Inconsistent behavioral anchors risk obscuring meaningful biological signals and hindering progress toward individualized care.

While some prior work has compared individual assessment scores or examined their psychometric properties [10], [8], these efforts have been limited in scope and sample size. Critically, no study has systematically evaluated both the semantic content and biological alignment of multiple ASD assessments across multiple datasets. To address this gap, we conducted the first large-scale comparative analysis of three widely used assessments: ADOS, ADI-R, and SRS. We quantified their semantic similarity using sentence-level embeddings generated by the BioSentVec language model, pre-trained on biomedical text corpora [11]. We then examined correlations among sub-scores across three publicly available ASD datasets comprising a total of N = 1 954 individuals—providing a robust sample size for cross-assessment comparison. Despite substantial semantic overlap in item content, the behavioral sub-scores were minimally correlated, with ADOS being the least aligned. Classifications based on standard thresholds exhibited only moderate agreement, ranging from 60–80% consistency across assessments. Finally, we evaluated associations between each assessment and commonly studied biological features derived from structural and functional MRI and genomic data. Our findings reveal substantial inconsistencies in biological correlates across datasets and modalities. Taken together, these results raise fundamental concerns about the reliability and validity of current behavioral assessments in autism and highlight the urgent need for clinical and diagnostic tools that are consistent, scalable, and biologically grounded.

## II. Results

### Semantic Overlap of Behavioral Assessments Does Not Translate to Score Reliability

Fig. 1 (top-left) shows the semantic similarity between the items that comprise each assessment sub-score, as measured using the inverse Wasserstein distance. As expected, items within the same assessment cluster strongly, producing a block-diagonal matrix structure. However, we also note crucial overlap across assessments. In particular, the item-level embeddings comprising the ADOS Social Affect score are highly similar to those of the ADI-A (Social Interaction) and ADI-B (Communication) sub-scores, the SRS Awareness, Cognition, and Communication sub-scores, and the SCQ. The ADI-A and ADI-B sub-scores show strong semantic similarity to the SRS Cognition and Communication sub-scores and to the SCQ.

**Figure 1.**
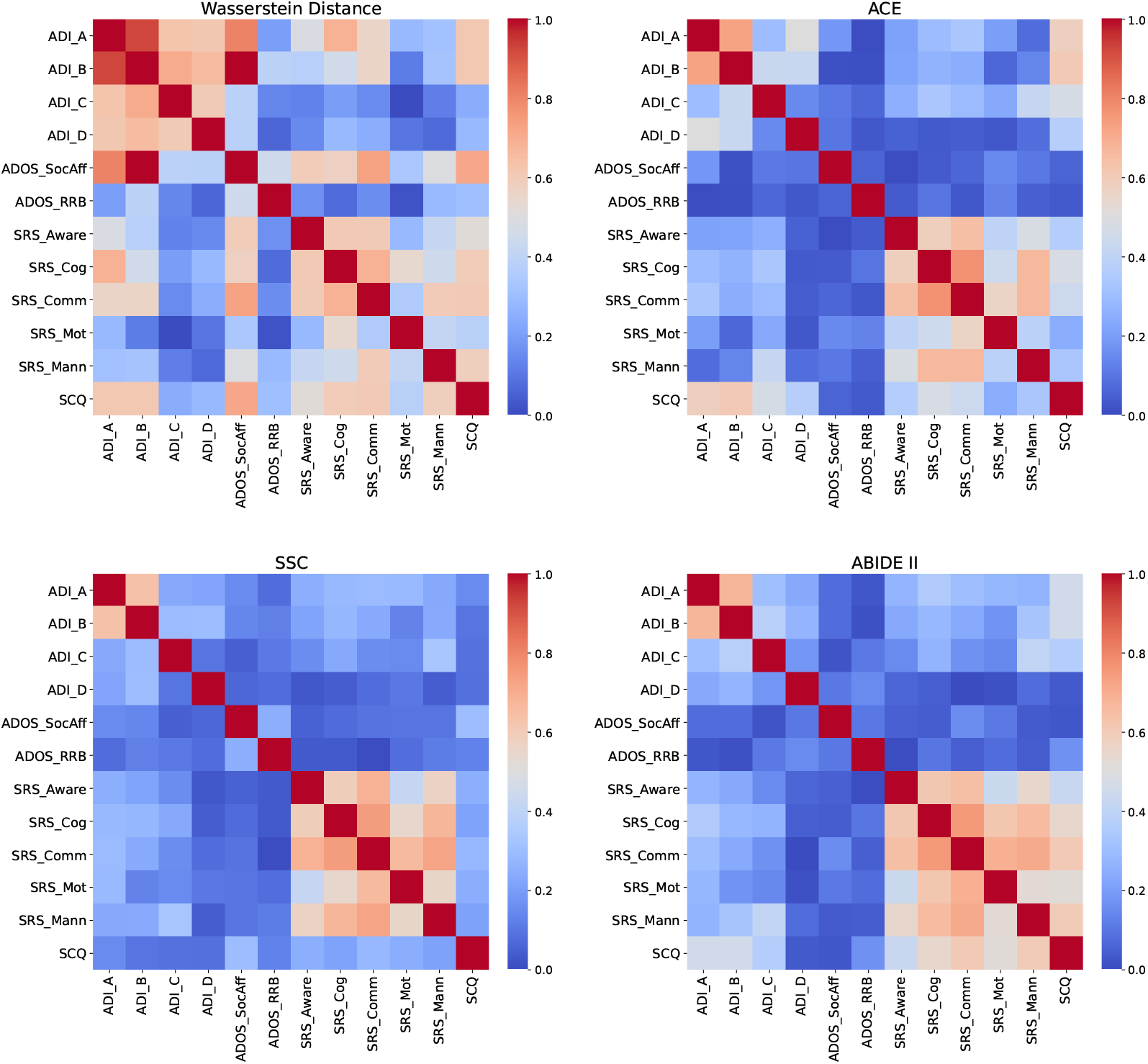
Comparison of item-level and sub-score associations across behavioral assessments. **Top Left:** Similarity of item-level sentence embeddings quantified using the inverse Wasserstein distance. **Top Right & Bottom:** Pearson correlation coefficient between the assessment sub-scores across participants in the ACE (Autism Center for Excellence), SSC (Simons Simplex Collection), and ABIDE II (Autism Brain Imaging Data Exchange II) datasets. Despite their high semantic similarity, the sub-scores are nearly uncorrelated across assessments. Sub-score abbreviations are described in Table S1 of the supplement.

The participant-level behavioral scores in Fig. 1 (top-right and bottom) point to a contrasting result, with significant correlations appearing only within the same assessment. The SRS demonstrated strong internal correlations among its own sub-scores (generally *r* > 0.5), which is consistent with the reported redundancy of the questionnaire [12]. However, the SRS exhibited little association with other tools. The ADOS Social Affect sub-score showed the weakest external alignment, with *r* < 0.2 in most cases. The ADI-A and ADI-B sub-scores were strongly correlated with each other, but the ADI-R as a whole did not meaningfully correlate with other measures aside from a moderate correlation with SCQ in the ACE dataset. Taken together, these results show that behavioral assessments, despite targeting overlapping constructs, yield inconsistent phenotypic profiles at the participant level.

To further quantify the discrepancies in Fig. 1, we measured the alignment between semantic similarity (inverse Wasserstein distance) and participant-level correlations of the behavioral sub-scores. Formally, let **w**_*i*_ and **a**_*i*_ denote the *i*^*th*^ rows of the Wasserstein matrix and a correlation matrix (excluding the diagonal). Using the linear model **a**_*i*_ = *β*_*i*_ **w**_*i*_, we report alignment coefficients *β*_*i*_ for each sub-score and dataset in Table I. Across datasets, the SRS showed the strongest alignment, largely driven by its high internal correlations. The ADI-R and SCQ demonstrated moderate but variable alignment (ADI-R: ∈ *β* [0.235, 0.578]; SCQ: *β* ∈ [0.353, 0.697]), while the ADOS was generally the least aligned (*β* ∈ [− 0.014, 0.417]). Overall, these results confirm that high semantic overlap does not translate into consistent behavioral measurements, with ADOS standing out as the most inconsistent across datasets.

### Data-Driven Patient Stratifications are Inconsistent Across Behavioral Assessments

To capture potential nonlinear relationships between assessments, we evaluated whether clusters derived from behavioral sub-scores were consistently reproduced across instruments. We applied *k*-means clustering to the sub-scores of each assessment separately (ADI-R, ADOS, SRS) and quantified the overlap in participant groupings across them. The clustering was repeated 100 times with random initializations, yielding interquartile ranges that confirmed reproducibility. The analyses were conducted separately for each dataset.

Fig. 2 reports the percentage overlap between participant groups generated for each assessment with *k* = 2, 3, 4, 5 clusters. Overall, clustering participants by behavioral subscores produced inconsistent groupings. Across datasets, the mean overlap was 60 *±* 5% for two clusters, with overlap declining as the number of clusters increased. The ADI-R and SRS clusters showed the highest agreement, consistent across datasets, whereas the ADI-R and ADOS clusters showed the lowest. Notably, both ADI-R and SRS are completed by caregivers, while ADOS is the only tool based on direct clinical observation. These results highlight a systematic divide between caregiver-reported and clinician-observed measures in shaping patient stratifications. Importantly, such inconsistencies pose a barrier to personalized care.

**Figure 2.**
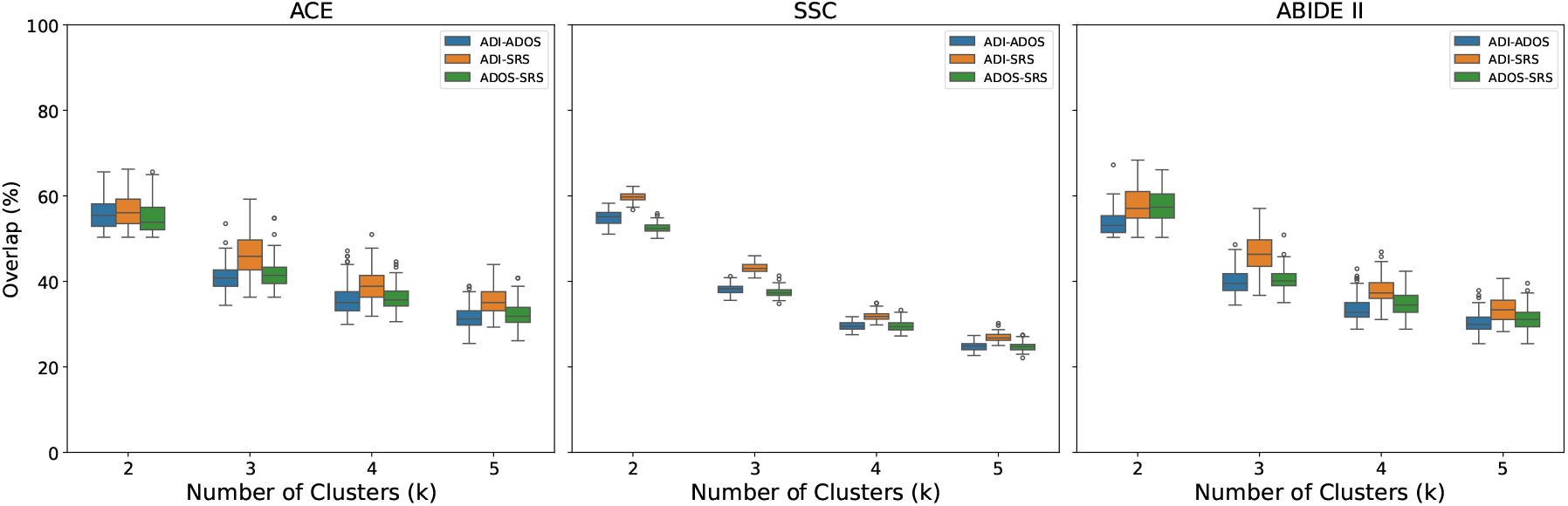
Consistency of participant stratifications across behavioral assessments. Percentage overlap between clusters derived from ADI-R, ADOS, and SRS sub-scores for *k* = 2, 3, 4, 5 clusters in the ACE, SSC, and ABIDE II datasets. While caregiver-reported assessments (ADI-R, SRS) show moderate overlap, clinician-administered ADOS clusters diverge substantially, highlighting inconsistencies in data-driven patient stratifications across assessments.

### Moderate Diagnostic Agreement Across ADOS and ADI-R

Given the weak correlations between ADOS and ADI-R sub-scores, we next evaluated whether these inconsistencies extend to their diagnostic classifications. For each dataset, participants were divided into “diagnosed” and “non-diagnosed” groups by thresholding the behavioral scores of each instrument (*score ≥ threshold* for diagnosis). Analyses were conducted separately on each dataset.

Fig. 3 shows the percentage of participants consistently classified between ADOS and ADI-R when thresholding their total raw scores on each assessment. Because no universally accepted thresholds exist, we included a range of values. Across datasets, diagnostic alignment was moderate, with agreements ranging from 65-80%. We note that the agreement is highly variable as the thresholds are varied, which raises concerns about the sensitivity of these instruments to minor scoring differences by the evaluators.

**Figure 3.**
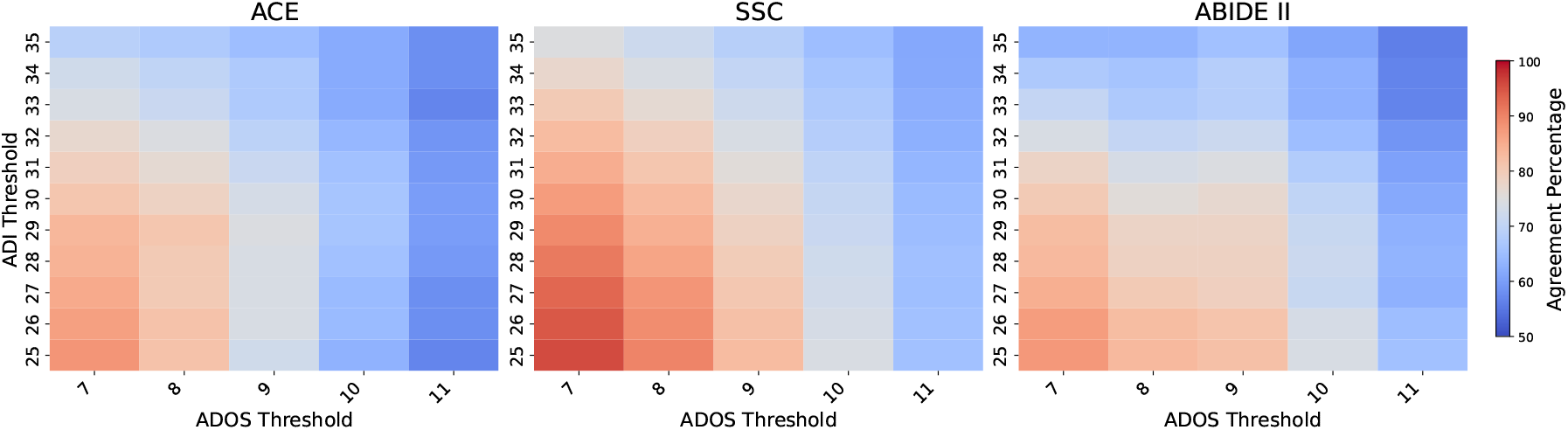
Diagnostic alignment between ADOS and ADI-R. Heatmaps show the percentage of consistently classified participants as a function of cutoff thresholds for each instrument. Agreement is highlighted for the commonly used threshold pair (ADOS: 9, ADI-R: 30), which yields only moderate alignment.

Even when selecting a specific common diagnostic threshold for the ADI-R sub-scores (ADI-A *≥* 10, ADI-B *≥* 7, ADI-C *≥* 3) and the ADOS (Total *≥* 9) confirms the trends in Fig. 3. All three datasets have moderate agreement across datasets (ACE: 67.52%; SSC: 78.95%; ABIDE II: 76.27%) with low Cohen’s *κ* values (ACE: *κ* = 0.076; SSC: *κ* = 0.031; ABIDE II: *κ* = 0.11), indicating only slight agreement beyond chance. Changing the thresholds for either test reduced alignment, suggesting that a high score on one instrument does not reliably map onto a higher score on the other.

Taken together, our findings demonstrate that even when applying standard diagnostic thresholds in high-quality research datasets, ADOS and ADI-R yield only moderate agreement. Thus, inconsistencies between the instruments can undermine reliable phenotyping in ASD research.

### Behavioral Sub-Scores Show Divergent Biomarker Associations Across Datasets and Modalities

Building on the inconsistencies observed at the behavioral level, we next asked whether these assessments show consistent associations with biomarkers from three complementary modalities: structural MRI, resting-state functional MRI, and genomic data. These modalities capture anatomical, functional, and genetic dimensions of ASD neurobiology. We focused on high-level biomarkers repeatedly implicated in the ASD literature, rather than an exhaustive search that would exacerbate the multiple comparison problem. This framework allowed us to test whether consistent biological associations emerge across datasets and modalities, or whether the variability in behavioral scores extends into their biological correlates.

For the genomic data, we compared polygenic risk score (PRS) distributions for ASD between participants with “low” and “high” behavioral scores, split at the dataset median. Analyses were performed on the ACE and SSC datasets, as ABIDE II does not provide genomic data. Fig. 4 shows the PRS distributions based on total raw scores for ADOS, ADI-R, and SRS. A Welch’s two-sample *t*-test with unequal variances revealed significant differences in PRS between groups for the ADI-R in the SSC dataset (ACE: *t* = 1.81, *p* = 0.14; SSC: *t* = 2.64, *p* = 0.02), whereas neither the ADOS nor the SRS showed significant group effects. These results suggest that genetic associations with ASD vary depending on the behavioral assessment used, underscoring the lack of consistent alignment with common genetic variation.

**Figure 4.**
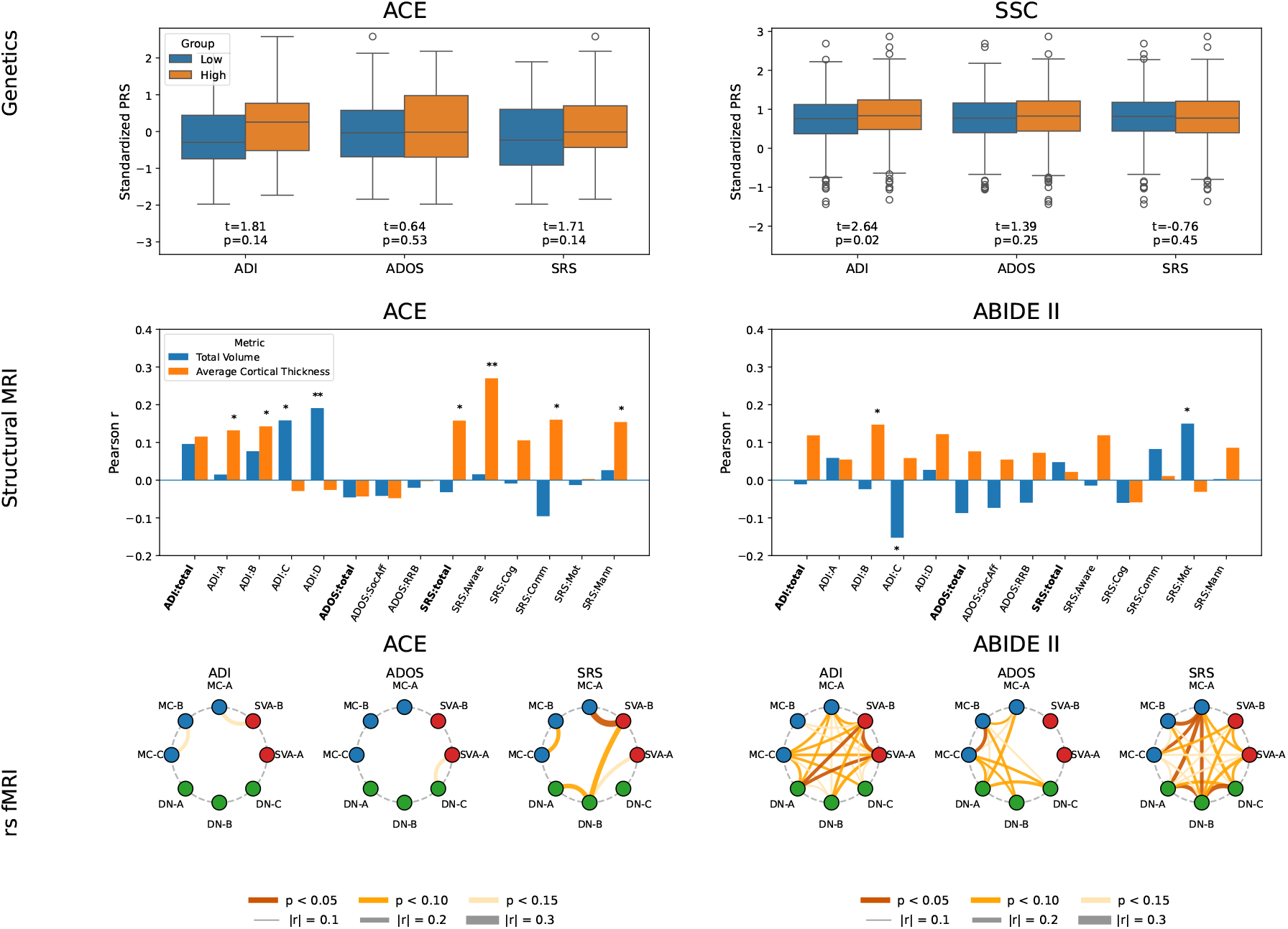
**Top (Genetics):** Polygenic risk score (PRS) associations with behavioral assessments. Distributions of ASD PRS for participants with “low” and “high” behavioral scores (median split) in ACE (left column) and SSC (right column). Significant group differences emerged only for the ADI-R, while ADOS and SRS showed no clear separation. **Middle (Structural MRI):** Pearson correlation for total cortical volume (blue) and average cortical thickness (orange) against total scores and sub-scores for ADI-R, ADOS, and SRS in ACE (left column) and ABIDE II (right column). Double asterisks indicate statistical significance (*p* < 0.05), while single asterisks mark strong but non-significant associations (*p* < 0.15). **Bottom (rs-fMRI):** Network-level associations with ADI-R, ADOS, and SRS in ACE (left column) and ABIDE II (right column). Brain regions arranged in a circle and color-coded by network family. Connections represent significant associations, with line color indicating statistical significance (*p* < 0.05, *p* < 0.10, *p* < 0.15) and line thickness reflecting inter-network functional connectivity strength.

For structural MRI data, we correlated behavioral subscores and raw total scores with two imaging metrics commonly linked to ASD: total brain volume and average cortical thickness. Fig. 4 reports the imaging–behavior correlations for ACE and ABIDE II. Double asterisks indicate statistical significance (*p* < 0.05), while single asterisks mark strong but non-significant associations (*p* < 0.15). Statistically significant correlations were observed between the SRS social awareness score and average cortical thickness in ACE. Significant results were also seen between total brain volume and the ADI-D score. However, these result did not translate to ABIDE II. Some strong associations were observed for both the ADI and SRS, and none were observed for ADOS. These findings suggest that structural MRI-behavioral associations are dataset-dependent and fail to show robust, cross-cohort consistency.

For rs-fMRI data, we computed functional connectivity between eight Yeo-17 atlas networks [13] previously linked to ASD (Salience/Ventral Attention A and B; Motor Control A, B, and C; Default Mode Networks A, B, and C) [14]. Each of the 28 unique connectivity values was correlated with the total raw scores of the ADI-R, ADOS, and SRS. Fig. 4 summarizes these results. In the ACE dataset, only one SRS association reached significance at *p* < 0.05, being between the Motor Control A and Salience/Ventral Attention Network B networks. Other strong but non-significant trends at *p* < 0.10 and *p* < 0.15 were observed for all three tests, with the only overlapped strong connectivity being between the ADI and the SRS in Motor Control regions.

In contrast, ABIDE II showed significant associations for all three behavioral assessments, yet there was minimal overlap among the identified connections. Specifically, connectivity between the Salience/Ventral Attention networks and Default Mode Network A was correlated with ADI-R scores; ADOS scores were significantly associated only with Motor Control B and C; and SRS scores showed the strongest associations with Default Mode and Motor Control networks. These findings highlight both the lack of cross-cohort consistency and the limited agreement between phenotype measures in their neural correlates, underscoring how variability across behavioral instruments complicates the search for reproducible neuroimaging biomarkers in ASD.

## III. Discussion

In this study, we conducted the first large-scale, multidataset comparison of widely used autism behavioral assessments and their biological correlates. Despite substantial semantic overlap among assessment items, we found minimal correlations between behavioral scores, only moderate diagnostic agreement between the gold-standard ADOS and ADI-R instruments, and divergent associations with neuroimaging and genetic biomarkers. After decades of research, no stable biomarkers for autism have emerged, and progress toward robust therapeutics remains limited. Our findings suggest that one underappreciated factor is the inconsistency of commonly used behavioral assessments, which—based on all evidence presented here—do not provide a uniform measure of autistic traits. These inconsistencies undermine efforts to define reproducible patient subtypes and complicate the search for meaningful biological markers or treatment targets.

Clinically, the ADOS, ADI-R, SRS, and SCQ each provide valuable but distinct vantage points on autistic traits–direct observation, developmental history, dimensional variation, and rapid screening, respectively. Yet this very diversity creates a troubling inconsistency: patients may be classified differently depending on which tool is used. In particular, the ADOS relies on clinician notes which are then reduced to numerical scores, which may obscure nuances that emerge during observation. Likewise, variations in the participant age at the time of assessment may influence the results. In sum, when instruments that ostensibly measure the same condition yield discordant outcomes, the reliability of both clinical decisionmaking and biomarker discovery is undermined.

Ours is not the only study that calls into question the reliability of current behavioral assessments. Frigaux et al. reported that even gold-standard tools such as the ADOS and ADI-R can fail in complex situations, revealing diagnostic challenges for infants and adults [15]. Dukevot et al. evaluated the screening accuracy of the SRS against the ADOS and showed that different informants provide unique, and sometimes conflicting, information for ASD assessment [16]. Boosalis identified multiple factors driving discrepancies between the ADOS and SRS, with low correlation coefficients between the two tests [10]. Sappok et al. examined the specificity and sensitivity of the ADOS and ADI-R in populations with intellectual disabilities, finding divergent results and recommending adjustments to improve feasibility and specificity [17]. Recent work has also highlighted there are multiple distinct manifestations of autism that may require different diagnostic criterion [18], [19] Building on these concerns, our study leverages multiple large, publicly available datasets to probe fine-grained clinical phenotypes and their biological correlates—revealing that even semantically similar assessments show minimal alignment with one another and no consistent relationship to neuroimaging or genetic markers.

One hypothesis for the inconsistent clinical phenotyping is that the scores reflect the subjective biases of the evaluator as much—or more—than the child’s underlying behavior. For example, both ADI and SRS rely on parent observations and show slightly higher correlations with each other in Fig. 1 than with ADOS, which is evaluated by a clinician. Likewise, participant stratifications in Fig. 2 are consistently more similar between ADI and SRS across datasets and cluster numbers. As further evidence, we examined a subset of SSC participants for whom the SRS was completed by both a parent and a teacher (N=1424). Fig. 5 shows that although the parent and teacher SRS total scores are correlated, their numerical values deviate by an average of 37.82%. A similar trend can be observed for the SRS sub-scores (Fig. S7 of the supplement). Taken together, these findings indicate that evaluator perspective can meaningfully shape the measured phenotype, which in turn can obscure genuine biomarker relationships and effective personalized interventions for ASD.

**Figure 5.**
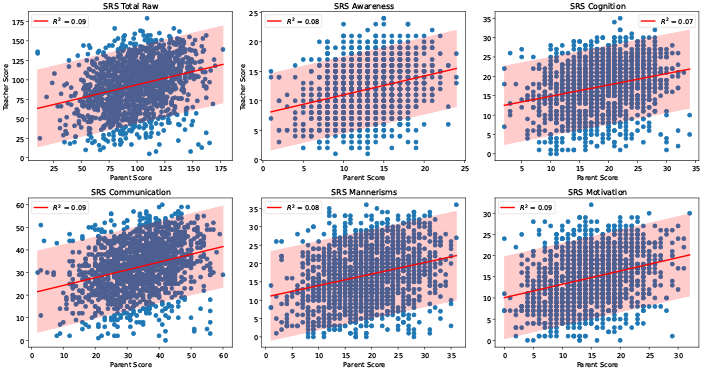
Comparison of the SRS total score and composite scores when the assessment is completed by a parent/caregiver and by a teacher. 90% prediction intervals are highlighted. The proportion of shared variance is low at *R*^2^ = 0.07 − 0.09.

Our analyses also show that no single behavioral assessment consistently outperforms the others in its alignment with biological markers. Associations with neuroimaging and genomic features varied by instrument and dataset, suggesting that each tool captures only a partial and context-dependent view of autistic traits. These discrepancies are consistent with prior reports that imaging correlates of ASD are often modest, inconsistent, or site-specific [20]. This lack of convergence highlights the need for behavioral instruments that are explicitly grounded in ASD neurobiology rather than expert consensus. Another key takeaway is the importance of validating both new and existing tools across larger, more diverse, and less curated cohorts to ensure that observed associations generalize beyond well-resourced research settings. Finally, integrating complementary assessments or combining behavioral data with objective measurements, such as eye-tracking or speech, may help to stabilize clinical phenotyping and provide a more reproducible anchor for biomarker discovery.

Overall, our results underscore the need for reorganization of and innovation in behavioral phenotyping in autism reserach. No single assessment consistently aligns with biological markers across datasets, suggesting that current instruments capture only partial and context-dependent representations of autistic traits. Future progress will require integrative approaches that combine information from multiple assessments and quantitative behavioral measures, and that explicitly link phenotypes to neurobiological mechanisms. Computational methods, including semantic modeling and multimodal data integration, offer a principled path toward harmonizing existing tools and guiding the development of next-generation measures that are scalable, reproducible, and biologically grounded. Advancing such frameworks will be essential for improving biomarker discovery, enabling robust stratification in clinical trials, and translating neurobiological insight into more precise and effective interventions.

Rather than diminishing the utility of behavioral assessment in autism. the inconsistencies identified here highlight a critical opportunity to refine how behavioral phenotypes are defined and operationalized. Existing instruments encode complementary clinical perspectives on autistic traits, shaped by differences in informant, context, and developmental focus. By systematically quantifying both their semantic overlap and empirical divergence, this study establishes a framework for understanding how measurement choices influence phenotypic definition and downstream biological inference. Such clarity is essential for harmonizing cohorts, improving reproducibility, and strengthening the linkage between behavior and neurobiology. Accordingly, our findings motivate a shift from reliance on single instruments toward integrative, biologically-informed phenotyping strategies that can better support translational autism research.

Our study is not without limitations. First, our analyses drew from curated research cohorts with high ASD likelihood and carefully collected data, which make our results *optimistic* compared to real-world clinical settings. In everyday practice, where resources and training vary widely, the inconsistencies we observed may be even greater. Second, autism diagnosis remains a bespoke process, guided by clinician observation, as informed by behavioral assessments. Varying emphasis on particular instruments, such as one clinician relying more heavily on the ADI-R while another prioritizes ADOS, can further distort phenotypic consistency across sites and studies. Finally, the neuroimaging and genetics features used in this study represent only a subset of possible biomarkers and may have missed subtle associations. In addition, some of the biomarker analyses were underpowered, which underscores the need for larger, harmonized datasets for ASD.

Future work should expand biomarker analyses to include additional imaging modalities, genomic measures, and longitudinal designs to test the stability of behavioral–biological relationships over time. To ensure robustness, candidate biomarkers should be validated across independent datasets and replicated in multiple studies before being advanced as potential diagnostic or therapeutic targets. Progress will also depend on greater multi-site collaboration and open data sharing to overcome underpowered analyses and site-specific effects. Parallel efforts should focus on developing improved behavioral instruments that more reliably capture the heterogeneity of ASD. Here, our strategy of systematically analyzing and comparing item semantics across assessments can assist in instrument harmonization for big data, precision phenotyping, and the creation of shorter, smarter screen measures. In sum, improving the reliability and biological validity of behavioral assessments is essential for establishing stable clinical phenotypes that can drive biomarker discovery and guide personalized care in autism.

## IV. Materials and Methods

### Datasets and Participant Selection

Analyses were conducted on three large, publicly available datasets. The ACE (Autism Center for Excellence) dataset was collected through a multi-site consortium led by the University of Virginia (R01 MH100028; PI: KAP). The network included five data-collection sites and a centralized data-coordinating center. The SSC (Simons Simplex Collection) [21] was compiled across 12 research clinics and aggregates genomic and phenotypic data from “simplex” families with a single diagnosed child. Participants were drawn from the imputed dataset available in SSC [22]. ABIDE II (Autism Brain Imaging Data Exchange) [23] is a multimodal neuroimaging repository that provides detailed phenotypic information. Data collection for ACE was conducted under Institutional Review Board approval at all participating sites. Further IRB details are provided in the supplement. The SSC and ABIDE II datasets are publicly available resources and were accessed in accordance with their respective data use agreements.

Participants were included in this study if complete data were available for all three behavioral assessments, ADOS Schedule 2, ADI-R, and SRS, along with the composite sub-scores for each instrument (ADOS: Social Affect and Restricted & Repetitive Behaviors; ADI-R: sub-scores A–D; SRS: Social Awareness, Social Cognition, Social Communication, Social Motivation & Mannerisms). This criterion biases the sample toward individuals suspected of ASD.

The SCQ was not available for all participants with complete ADOS Schedule 2, ADI-R, and SRS data. Accordingly, analyses involving the SCQ in Fig. 1 and Table I were restricted to the subset of participants with all four instruments. Likewise, not all participants had neuroimaging or genomic data available, and analyses involving those modalities were conducted on reduced sub-samples.

**Table I:** Alignment between semantic similarity (inverse Wasserstein distance) and behavioral correlations for each sub-score and dataset. The SRS showed the strongest alignment, while the Adi-R and SCQ had moderate but variable alignment. ADOS was the least aligned, underscoring its inconsistency with other assessments.

| Sub-Score | ACE | SSC | ABIDE II |
| --- | --- | --- | --- |
| ADI-A (Social Interaction) | 0.578 | 0.447 | 0.518 |
| ADI-B (Communication) | 0.498 | 0.408 | 0.462 |
| ADI-C (Early Development) | 0.546 | 0.379 | 0.512 |
| ADI-D (Impairment) | 0.527 | 0.235 | 0.303 |
| ADOS SocAff (Social Affect) | 0.057 | 0.200 | 0.107 |
| ADOS RRB (Restrict. & Repet. Behav.) | 0.065 | 0.417 | -0.014 |
| SRS Aware (Social Awareness) | 0.705 | 0.738 | 0.780 |
| SRS Cog (Social Cognition) | 0.737 | 0.716 | 0.813 |
| SRS Comm (Social Communication) | 0.746 | 0.722 | 0.805 |
| SRS Mot (Social Motivation) | 0.894 | 1.057 | 1.212 |
| SRS Mann (Mannerisms) | 0.740 | 0.887 | 0.982 |
| SCQ (Total Score) | 0.683 | 0.353 | 0.697 |

The ACE dataset included 157 participants, with 154 of them having the SCQ, 138 with structural MRI, 95 with functional MRI, and 91 with genomics data. The SSC dataset had a sample size of 1 620 participants. Out of these, 704 included the SCQ and 1 260 had genomics data. The ABIDE II dataset comprised of 177 participants. 96 had the SCQ, 135 had structural MRI data, and 177 had functional MRI data.

Finally, ABIDE II contains an additional N = 123 participants who were administered the ADOS-Generic [24] evaluation in addition to the ADI-R and SRS. Because prior work has shown that ADOS Schedule 2 and ADOS-Generic are not interchangeable [25], these participants were excluded from our primary study. Parallel analyses using the ADOS-Generic sample are provided in Fig. S3 of the supplement.

### Processing and Analyses of Behavioral Data

Semantic similarity between assessments was computed in three steps. First, individual assessment items (e.g., questions or observational criteria) were encoded using the BioSentVec large-language model [11], which is trained on biomedical texts. Second, item-level embeddings were grouped according to their sub-scores. Third, semantic relationships between subscores were quantified using the Wasserstein distance [26], which computes the optimal transport cost between two distributions. A smaller distance indicates greater semantic similarity between two sub-scores. Following standard practice in natural language processing, the Wasserstein distance was computed using a geodesic metric on the sphere [27], [28]. Distances were then normalized and inverted so that larger values indicate greater semantic similarity. We consider other large-language models in Fig. S1 of the supplement.

Independent of the semantic analysis, we computed the Pearson correlation coefficients between sub-scores of the ADOS, ADI-R, and SRS using participant behavioral data. For comparison, we also correlated each sub-score with the SCQ, which serves as a screening rather than a diagnostic tool; only participants with SCQ data were included in this analysis. We report the correlation magnitudes to examine association strength rather than direction. Raw scores were used for all four assessments. While some ASD studies rely on t-scores, this amounts to a linear transformation and would not change the correlation coefficients reported in this study. In addition, t-scores are sample-dependent, which reduces comparability across datasets. All analyses were conducted in Python 3.12.7 using NumPy (v1.26.4) and Pandas (v2.2.3).

We examined the consistency of patient stratifications by clustering participants separately on ADOS, ADI-R, and SRS sub-scores. Clustering was performed with the *k*-means algorithm using random initialization, with *k* = 2, 3, 4, 5. To account for algorithmic and sample variability, we generated 100 bootstrap subsets of the data and repeated clustering on each. Cluster overlap between instruments was quantified as the percentage of participants assigned to the same cluster after optimal label matching with the Hungarian algorithm. Overlap was computed for each bootstrap replicate, yielding a distribution of stratification congruency for each *k*.

We quantified diagnostic agreement between ADOS and ADI-R by classifying participants as “diagnosed” or “not diagnosed” based on specified thresholds. Following convention, ADOS was thresholded on the total score. For ADI-R, we examined two cases: thresholding the total score and thresholding individual sub-scores; in the latter case, all subscores were required to exceed their thresholds for diagnosis. Alignment was measured as the percentage of participants receiving the same diagnostic label on both tests, and Cohen’s *κ* was computed to estimate agreement beyond chance. To assess how the results vary across cutoff criteria, we conducted a grid search over common threshold ranges for each instrument and display the results as a heatmap.

### Genomics Data Pre-processing and Behavioral Association

Participants in SSC were genotyped in batches on Illumina Omni2.5, 1Mv3, or 1Mv1 arrays, and those in ACE on the Illumina HumanOmni 2.5M BeadChip. Quality control was performed separately for each SSC batch and for ACE to ensure comparability across platforms. Individuals appearing in both cohorts were removed from SSC to preserve sample independence. Analyses were restricted to a European subpopulation, defined by projecting participant genotypes onto the first three principal components of the 1 000 Genomes reference panel [29] and retaining individuals within 10 standard deviations of the European centroid. Only autosomal SNPs with call rate *≥* 99%, Hardy–Weinberg equilibrium *p* > 1 *×* 10^*−*6^, minor allele frequency *≥* 0.01, and hardcall threshold *≤* 0.1 were retained. Individuals with > 0.05 genotype missingness were excluded. To harmonize SSC and ACE, only SNPs present in both cohorts were retained. Linkage disequilibrium (LD) was estimated using the European subset of the 1 000 Genomes Project, with clumping at *R*^2^ < 0.1. Related individuals were excluded at 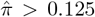 in PLINK [30].

We computed an ASD polygenic risk score (PRS) using GWAS summary statistics from iPSYCH [31] and SPARK [32], which do not include SSC or ACE participants. Following Weiner et al. [22], we applied a significance threshold *p ≤* 0.1, which explained the greatest case-pseudocontrol variance in SSC. The final PRS computation included 44 102 single nucelotide polymorphisms (SNPs) and was computed for each participant in SSC and ACE.

To examine the genomics-behavioral association, we first divided the participants into “low” and “high” groups based on the median value for each behavioral score. For each score and dataset, the PRS between the two groups was compared using Welch’s two-sample t-test with unequal variances. Benjamini-Hochberg’s false discovery rate procedure was applied for multiple comparisons.

### Structural MRI Data Pre-processing and Behavioral Association

MRI data for ACE were acquired on either a Siemens 3T Trio scanners with 12-channel head coils or a Siemens 3T Prisma scanners with 20-channel head coils using a standardized protocol adopted from the Human Connectome Project [33]. T1-weighted structural scans were preprocessed with sMRIPrep [34], which corrected for intensity inhomogeneity, performed skull stripping, segmented tissue types, and aligned images to standard space. FreeSurfer [35] was used to reconstruct the cortical surfaces for metric computation.

MRI data for ABIDE II was collected with varying scanners and acquisition protocols [23]. Given the larger sample, preprocessing used FastSurfer, a GPU-accelerated version of FreeSurfer that performs both the initial sMRIPrep preprocessing and the same cortical reconstruction steps. In rare cases where FastSurfer failed (N=7), standard FreeSurfer was used as an alternative. Participants whose data exhibited considerable artifacts after preprocessing were excluded.

We evaluate MRI-behavioral association based on two aggregate metrics, total brain volume and average cortical thickness across the brain, which are widely regarded as standard neuroanatomical biomarkers for ASD [36], [37], [38]. Pearson correlations were computed between each behavioral sub-score and MRI metric. Significance was determined at *p* < 0.05; we have also annotated associations with *p* < 0.15 for completeness.

### FMRI Data Pre-processing and Behavioral Association

Functional MRI data for ACE and ABIDE II were acquired using the scanners and head coils described above. TR was uniformly set to 2*s* for ACE but varied across ABIDE II sites. ACE fMRI data were preprocessed with fMRIPrep 23.0.2 [34], including head-motion correction, co-registration to the T1-weighted reference in MNI152 space, confound estimation, ICA-AROMA denoising, and resampling to volumetric and surface spaces. ABIDE II data were processed with fMRIPrep 25.1.4 using the same steps except ICA-AROMA was omitted.

The Yeo 17-network atlas [13] was used to parcellate the cortex into 17 networks. From here, eight networks were selected for analysis due to their reported ASD associations: Salience/Ventral Attention (#7, #8), Central Executive (#11–#13), and Default Mode (#14–#16) [14].

Functional connectivity between these eight networks was computed as the Pearson correlation coefficient between mean BOLD time series of all voxels within the respective network. This procedure yields 28 unique connectivity values. These connectivity measures were then correlated with ADOS, SRS, and ADI sub-scores to quantify the imaging-behavior associations.

## Supporting information

All supplemental information

## Acknowledgements

This work was supported by the National Institutes of Health awards R01HD108790 (Venkataraman), R01MH100028 (Pelphrey), and R01EB029977 (Caffo). This work was also supported by the Simons Foundation and the Simons Simplex Collection (13211.1, 15323.1; AV).

The authors would also like to thank Dr. Sayan Ghosal and Mr. Jeff Elibott for their assistance in preprocessing the genomics and ACE MRI data, respectively.

## Conflict of Interest

The authors declare no competing interests.

The Health Sciences Institutional Review Board of the University of Virginia gave ethical approval of research involving the ACE dataset. The Charles River Campus Institutional Review Board of Boston University waived ethical approval of research using the ABIDE II and SSC datasets.

## Data, Materials, and Code Availability

Python code used for data processing, analysis, and figure generation is publicly available at the following link: https://github.com/enrique-perezb/unreliable-clinical-phenotyping. The repository includes phenotypic data and summary-level neuroimaging and genetic features from ABIDE II, which is openly accessible without additional licensing.

Data from the ACE and SSC cohorts will be added pending approval from their respective consortia.

