## Supplementary material for "Behavioral Assessment Reliability in Clinical Phenotyping and Biomarker Research for Autism": All supplemental information

#### 1 Main Text Section: *Semantic Overlap of Behavioral Assessments Does Not Translate to Score Reliability*

##### 1.1 Nomenclature for Behavioral Assessment Sub-Scores

ADI-R, ADOS 2, and SRS include multiple sub-scores that capture distinct dimensions of ASD. Table 1 lists the sub-scores and corresponding abbreviations used throughout the main text.

##### 1.2 Different LLM Embedding Tools Maintain the Overall Conclusions

Our study uses the BioSentVec large language model (LLM) to generate sentence-level embeddings that we use to quantify the semantic relationships between the behavioral assessments.

We confirm the reliability of these semantic relationships across four popular LLMs: SentenceBERT [1], Word2Vec [2], BioSentVec [3], and BioWordVec [4]. SentenceBERT is a general-purpose model for sentence embeddings, whereas Word2Vec provides word-level embeddings that we averaged to obtain sentence-level representations. BioSentVec and BioWordVec are trained on biomedical corpora, producing sentence- and word-level embeddings, respectively.

Figure 1 compares the semantic similarity, as computed by the inverse Wasserstein distance between sentence embeddings. We observe that the embedding relationships are qualitatively similar across LLMs, including the high similarity between the ADOS Social Affect sub-score and the ADI-A and ADI-B subscores. We emphasize that the LLM embeddings are only used to construct the Wasserstein matrix in Fig. 1 of the main text to support the semantic similarities across assessments. The choice of LLM *does not influence any other result or conclusion of the paper*, including the inconsistencies between instruments, the mismatched phenotypic clusters, moderate diagnostic alignment, and biomarker associations.

##### 1.3 Subject-Level Data Support Inconsistencies in Autism Phenotyping

To further investigate the unreliable clinical phenotypes observed in this study, we plot subject-level associations between the total raw scores of each assessment in Fig. 2. As expected, we observe a low shared variance between all tests. Specifically, the shared variance between ADOS and ADI ranged from $R^2 = 0.01 - 0.03$ across datasets. The SRS and ADI had the highest shared variance, ranging from $R^2 = 0.10 - 0.13$ across datasets. Finally, the ADOS and SRS had the lowest shared variance at $R^2 < 0.01$. The 90% prediction intervals also highlight the lack of prediction capabilities from one test to another. In sum, these subject-level results further underscore our main conclusions of unreliability and a lack of precision phenotype across behavioral instruments.

##### 1.4 Substituting ADOS-G for ADOS-2 Does Not Improve Alignment with Other Measures

As noted in the main text, N=123 subjects in the ABIDE II dataset were administered ADOS-G instead of ADOS 2, along with both the ADI-R and SRS. These subjects were excluded from our study due to the reported differences between ADOS-G and ADOS 2. For completeness, Fig. 3 reports the behavioral correlations for these N=123 subjects when using ADOS-G in place of ADOS 2. Once again, we observe that the correlations between ADOS-G sub-scores and all other assessments remains low ($R < 0.3$). However, the ADOS-G showed significantly higher self-correlation between its sub-scores (Communication, Social Behavior, and Repetitive and Stereotyped Behavior) despite them quantifying distinct dimensions of ASD.

#### 2 Main Text Section: *Data-Driven Patient Stratifications are Inconsistent Across Behavioral Assessments*

In the main text, we compared the data-driven phenotypic clusters derived from ADI, ADOS and SRS on a pairwise basis. To further examine the inconsistencies, Fig. 4 reports the percentage overlap in the clusters when compared across all three instruments, simultaneously. These results reveal near-random overlap, indicating that different behavioral assessments produce inconsistent groupings. Across datasets, the mean overlap across the three behavioral instruments was $35 \pm 7\%$ for two clusters, with the percentage overlap decreasing as the number of clusters increased.

#### 3 Main Text Section: *Behavioral Sub-Scores Show Divergent Biomarker Associations Across Datasets and Modalities*

##### 3.1 No Significant Associations Between PRS and Behavioral Sub-Scores

As a supplement to the relationship between poly-genetic risk score (PRS) for ASD and the behavioral assessments, Fig. 5 reports the associations with each sub-score using the same procedure outlined in the main text. We do not observe any significant associations.

### 4 Main Text Section: *Discussion*

#### 4.1 SRS Scores By Parents and Teachers Exhibit Low Shared Variance

Using the SSC dataset, we examined the consistency in SRS sub-scores when the assessment is completed by a parent and by a teacher for the same subject. As expected, the results in Fig. 6 show a low shared variance between examiners, with $R^2$ ranging between 0.07–0.09 for all sub-scores.

We also analyze the SRS as a diagnostic tool, using a threshold of 75 on the total raw score to determine ASD ($>= 75$) versus neurotypical ($< 75$). Fig. 7 shows that 28.5% of participants have different binary ASD classifications when the test is conducted by a teacher and a parent. These results call into question the reliability and objectivity of the test, as the respondent appears to be a crucial aspect that significantly varies the results.

### 5 Main Text Section: *Materials and Methods*

#### 5.1 Participant Sample Sizes

Table 2 shows the participant sample sizes in table format.

#### 5.2 IRB Approval Information for ACE

Data from Wave 1 of the original ACE GENDAAR project were collected across four sites: Yale University, Harvard University/Boston Children's Hospital, the University of California, Los Angeles, and the University of Washington/Seattle Children's Research Institute. All procedures were approved by the institutional review boards (IRBs) of participating sites and conducted in accordance with the Declaration of Helsinki. The study received IRB approvals as follows: Yale as lead institution (IRB: 1206010363; Pelphrey), UCLA (IRB: 10-000387-CR-00008; Bookheimer), Boston Children's Hospital (IRB: P00004852; Nelson), and with Seattle Children's Hospital (IRB: 00000277; Webb) as a reliant site to Yale. George Washington University (GWU) assumed the role of lead institution (IRB 031802; Pelphrey) and the ACE project is now governed as a single-site IRB at the University of Virginia (UVA; IRB-HSR-220423; Pelphrey).

The ACE GENDAAR Data Coordination Center (DCC) was originally governed by UCLA, subsequently by the University of Southern California (USC; HS-18-00467, HS-13-00668; Van Horn), and presently by UVA (IRB-HSR-22078; Van Horn).

De-linked phenotypic, neuroimaging, and genetics data from ACE GENDAAR Wave 1 has been shared with the NIMH Data Archive (NDA), beginning 11/23/2012, under study collection ID 2021.

| Acronym | Behavioral Assessment Sub-Score |
|---|---|
| ADI_A | ADI Social Interaction |
| ADI_B | ADI Communication |
| ADI_C | ADI Restricted, Repetitive and Stereotyped Patterns of Behavior |
| ADI_D | ADI Abnormality of Development Evident at or Before 36 Months |
| ADOS_SocAff | ADOS Social Affect |
| ADOS_RRB | ADOS Restricted and Repetitive Behaviors |
| SRS_Aware | SRS Social Awareness |
| SRS_Cog | SRS Social Cognition |
| SRS_Comm | SRS Social Communication |
| SRS_Mot | SRS Social Motivation |
| SRS_Mann | SRS Restricted Interests and Repetitive Mannerisms |

Table 1: Behavioral Assessment Sub-Scores with Full Names

Table 2: Sample sizes across datasets and analyses. Number of participants included from ACE, SSC, and ABIDE II for behavioral, neuroimaging, and genomic analyses. Sample sizes vary depending on the availability of each assessment and modality, with smaller subsets contributing to SCQ, neuroimaging, and genomic analyses.

| Dataset | Sample Size | w/SCQ | w/MRI | w/fMRI | w/Genomics |
|---|---|---|---|---|---|
| ACE | 157 | 154 | 138 | 94 | 91 |
| SSC | 1620 | 704 | — | — | 1260 |
| ABIDE II | 177 | 96 | 135 | 177 | — |

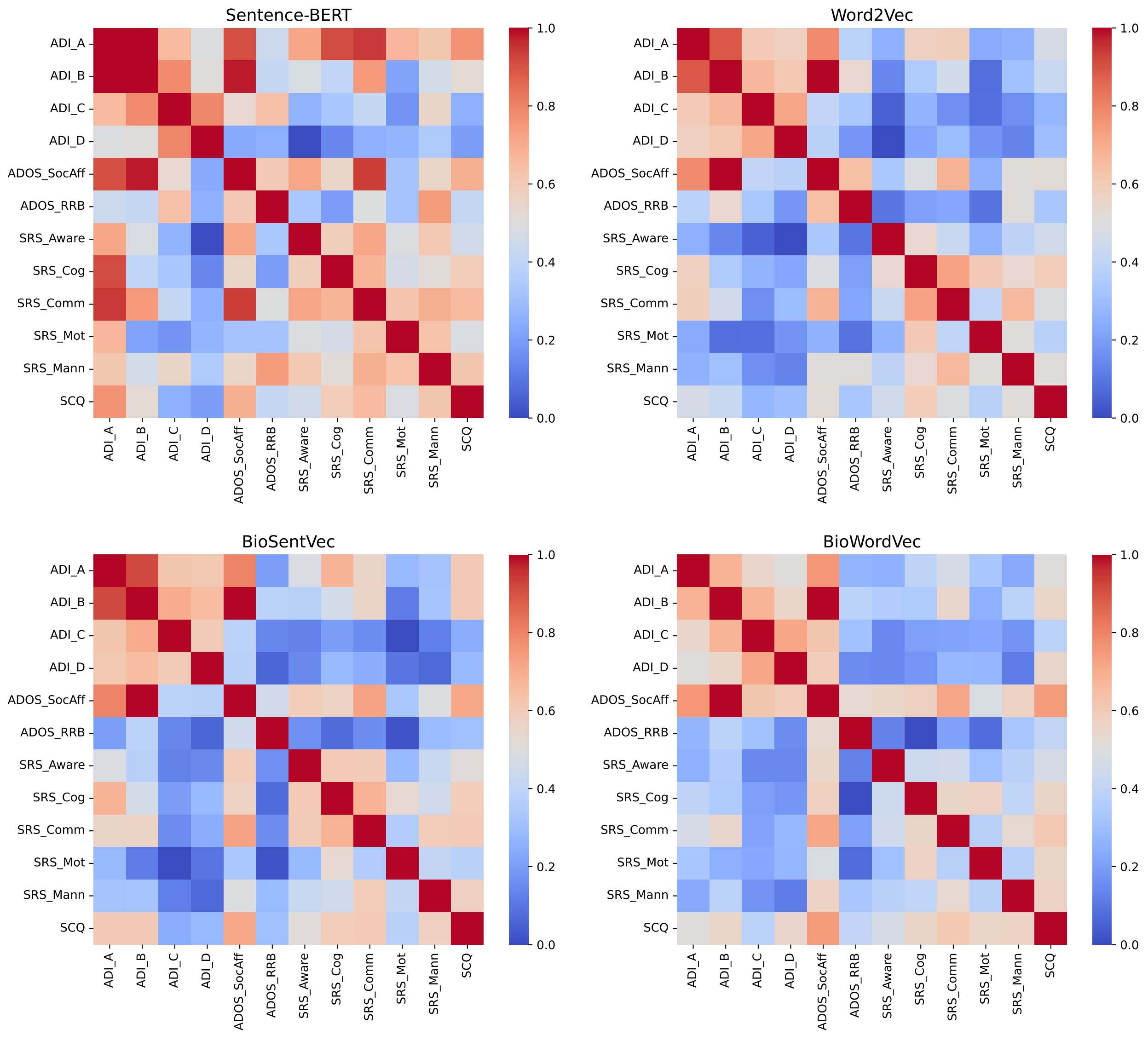


Figure 1: Comparison of embedding tools used to compute the inverse Wasserstein distance between sentence embeddings. Two general models (SentenceBERT, Word2Vec) and two biology-centric models (BioSentVec, BioWordVec) were used. Across models, embedding relationships remained similar.

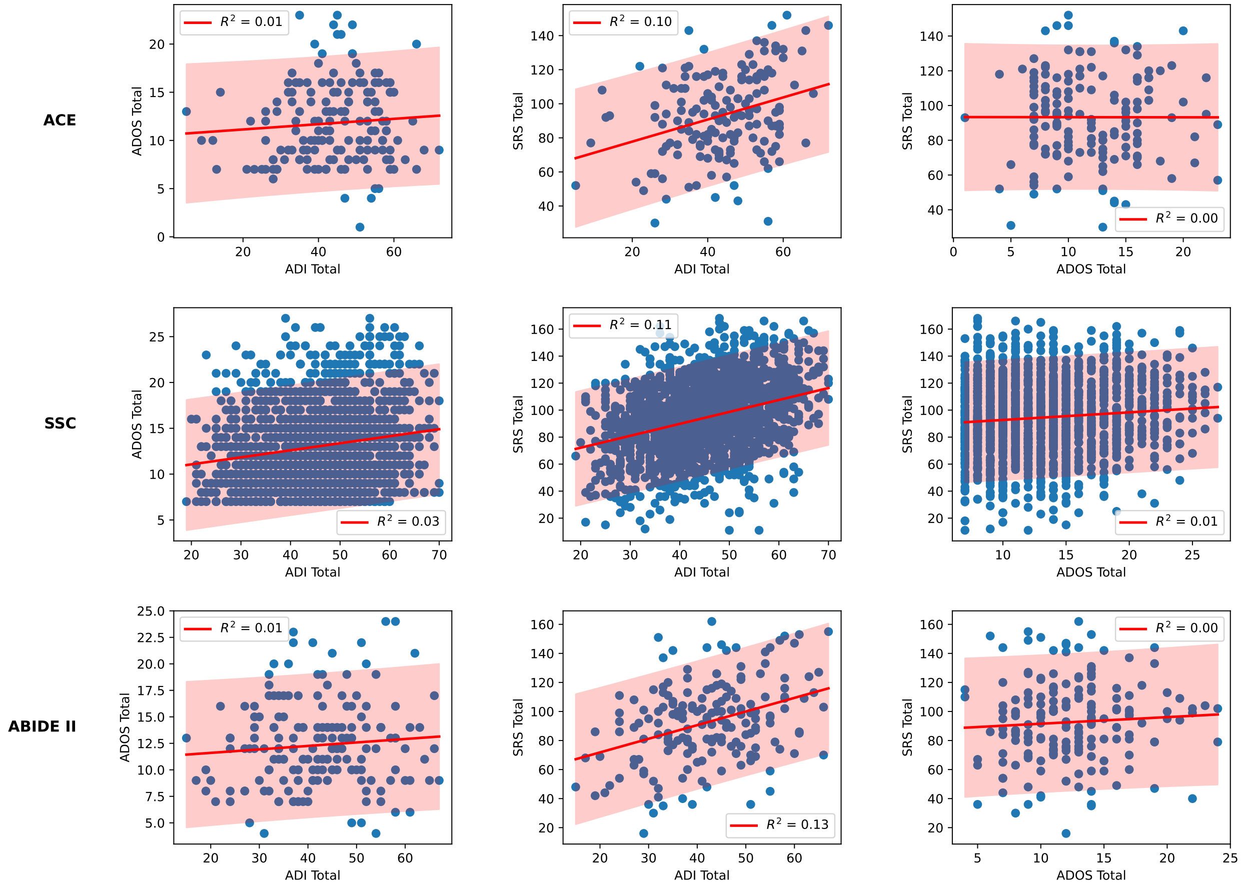


Figure 2: Pairwise comparison of total raw scores between the ADI, ADOS and SRS for all three datasets used (ACE, top; SSC, middle; ABIDE II, bottom). 90% prediction intervals are highlighted. The proportion of shared variance is low for all, with $R^2$ ranging from 0.00 to 0.13. The ADI and SRS appear to have the largest shared variance.

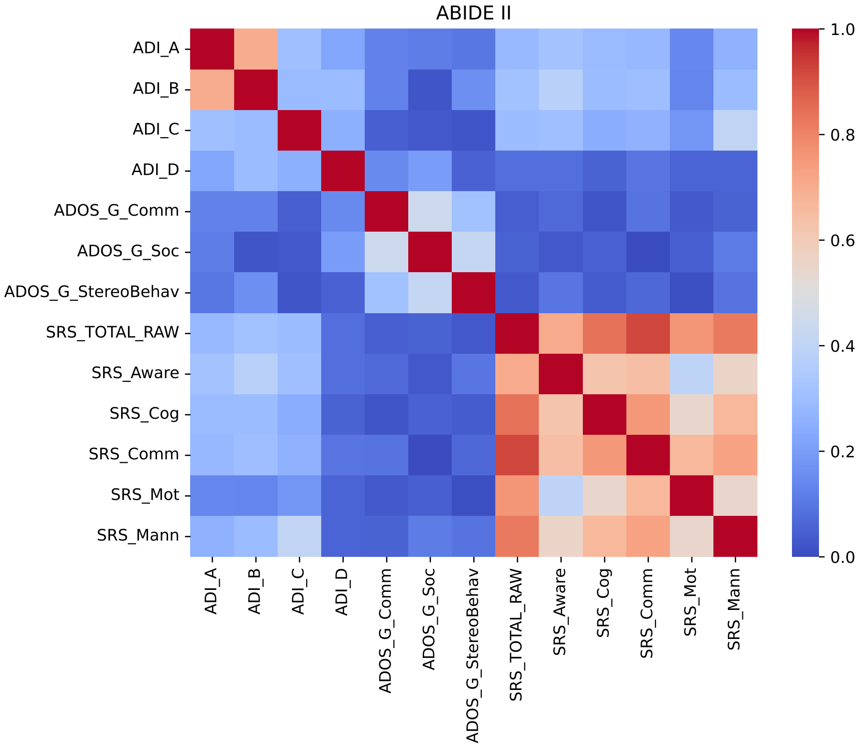


Figure 3: Comparison of item-level and sub-score associations across behavioral assessments for n=123 participants with the ADOS-G. Pearson correlation coefficient between the assessment sub-scores across participants in the ABIDE II dataset. The ADOS-G sub-scores are nearly uncorrelated across assessments.

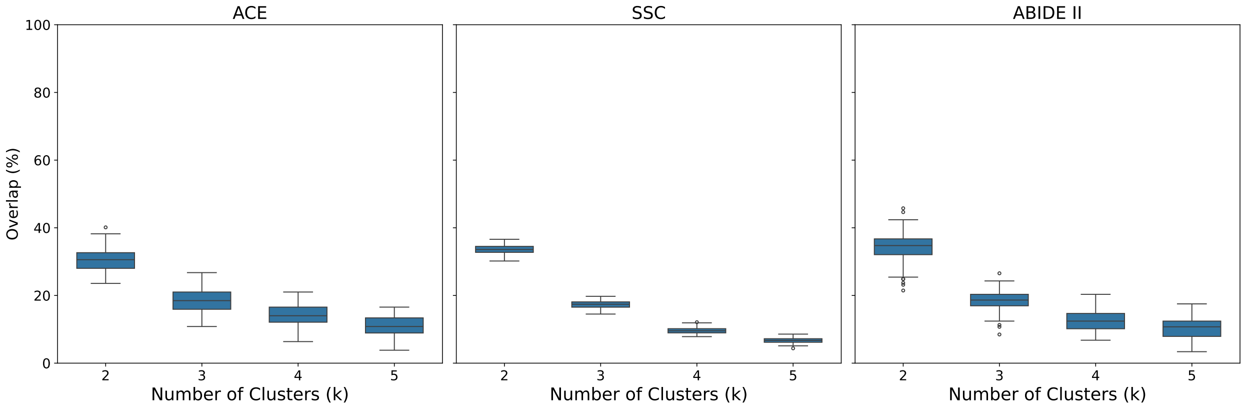


Figure 4: Consistency of participant stratifications across behavioral assessments. Percentage overlap between clusters derived from ADI-R, ADOS, and SRS sub-scores for $k = 2, 3, 4, 5$ clusters in the ACE, SSC, and ABIDE II datasets. Comparing the clusters between all three tests together shows near-random overlap percentage.

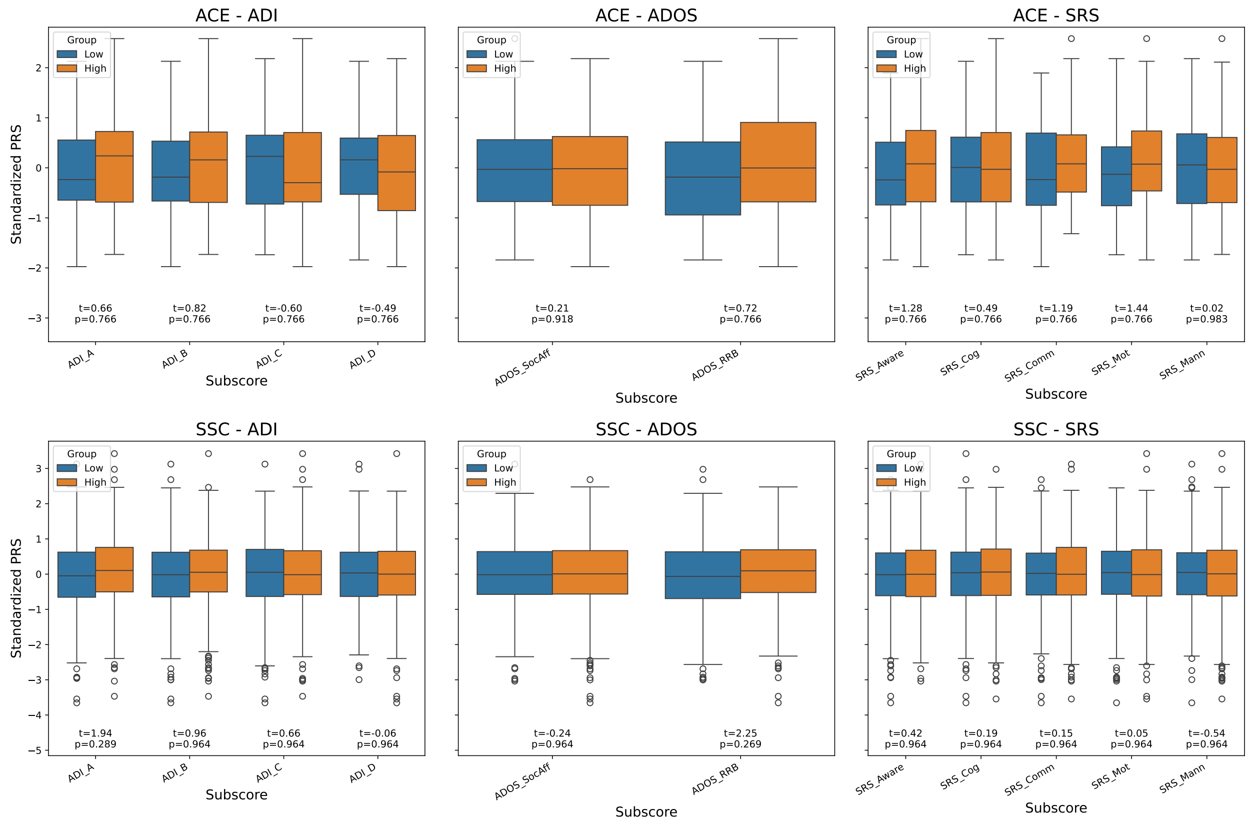


Figure 5: Polygenic Risk Score (PRS) associations with behavioral assessment sub-scores. Distributions of ASD PRS for participants with ”low” and ”high” behavioral scores (median split) in the ACE and SSC datasets. No significant differences emerged.

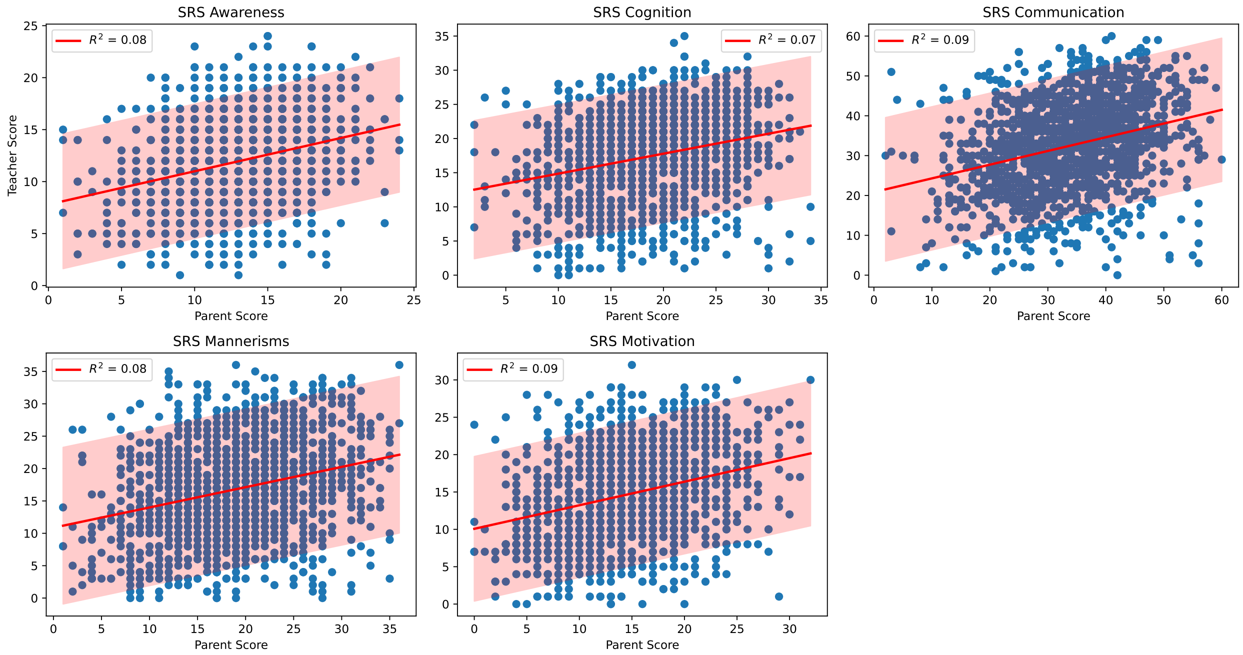


Figure 6: Comparison of the SRS sub-scores, when the assessment is completed by a parent/caregiver and by a teacher. 90% prediction intervals are highlighted. The $R^2$ is low for all, ranging between 0.07 and 0.09.

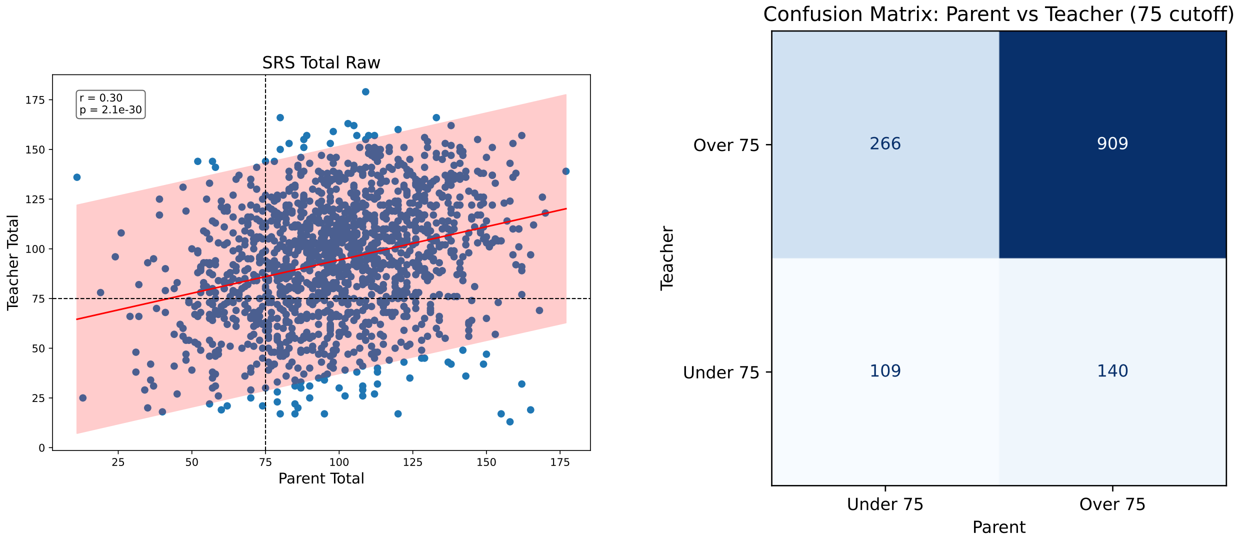


Figure 7: (Left) Comparison of SRS total raw scores from parent/caregiver and teacher assessments, with 90% prediction intervals and diagnostic threshold at 75. (Right) Confusion matrix of classifications based on these thresholds.

[2] Tomas Mikolov, Ilya Sutskever, Kai Chen, Greg S Corrado, and Jeff Dean. Distributed representations of words and phrases and their compositionality. In C.J. Burges, L. Bottou, M. Welling, Z. Ghahramani, and K.Q. Weinberger, editors, *Advances in Neural Information Processing Systems*, volume 26. Curran Associates, Inc., 2013.

[3] Qingyu Chen, Yifan Peng, and Zhiyong Iu. Biosentvec: creating sentence embeddings for biomedical texts. In *The Seventh IEEE International Conference on Healthcare Informatics*, 2019.

[4] Zhang Yijia, Qingyu Chen, Zhihao Yang, Hongfei Lin, and Zhiyong lu. Biowordvec, improving biomedical word embeddings with subword information and mesh. *Scientific Data*, 6, 2019.
